# Risk of Secondary Malignancies in Patients with prostate cancer: A Systematic Review and Meta-analysis

**DOI:** 10.1101/2020.03.31.20049098

**Authors:** Keyvan Heydari, Sahar Rismantab, Amir Shamshirian, Pouya Houshmand, Parisa Lotfi, Sajjad Rafati, Amir Aref, Ali Saravi, Danial Shamshirian, Behdad Zibaei, Reza Alizadeh-Navaei

**Affiliations:** Student Research Committee, School of Medicine, Mazandaran University of Medical Sciences, Sari, Iran; Gastrointestinal Cancer Research Center, Mazandaran University of Medical Sciences, Sari, Iran; Ramsar Campus, Mazandaran University of Medical Sciences, Ramsar, Iran; Department of Medical Laboratory Sciences, Student Research Committee, School of Allied Medical Sciences, Mazandaran University of Medical Sciences, Sari, Iran; Faculty of Veterinary Medicine, University of Tehran, Tehran, Iran; Belfer Center for Applied Cancer Science, Department of Medical Oncology, Dana-Farber Cancer Institute, Harvard Medical School, Boston, MA, USA; Student in Medicine, School of Medicine, Guilan University of Medical Sciences, Rasht, Iran; Chronic Respiratory Diseases Research Center, National Research Institute of Tuberculosis and Lung Diseases (NRITLD), Shahid Beheshti University of Medical Sciences, Tehran, Iran; Student in Medicine, School of Medicine, Gonabad University of Medical Sciences, Gonabad, Iran

**Keywords:** Prostate Tumor, Prostate Neoplasm, Secondary Malignancy, Meta-analysis

## Abstract

**IMPORTANCE:** Prostate cancer (PC) is the second most common cancer among males globally, however, the survival rate is favorable in most patients. In a small number of patients, who suffer from advanced or invasive cancer, various side effects such as secondary malignancies or treatment-related secondary malignancies (SMs) may be seen.

**OBJECTIVE:** To systematically asses the risk of secondary malignancies in patients with prostate cancer.

**DATA SOURCES:** We have searched for longitudinal studies through databases of Web of Science, Scopus and PubMed for all available data up to September 2019.

**STUDY SELECTION:** Studies with longitudinal design on prostate cancer patients that declared the results in SIR or those that the SIR could be calculated were eligible.

**DATA EXTRACTION AND SYNTHESIS:** The heterogeneity was evaluated using the I^2^ test. According to the results and in case of I^2^ ≥ 50%, the random effect model was used to combine the results. To identify the cause of heterogeneity in the studies, the analysis of sub-groups was performed based on the site of secondary malignancy, the treatment procedure, and duration of follow-up. Data were analyzed using STATA version 11.

**MAIN OUTCOMES AND MEASURES:** Overall SIR and based on treatment of prostate cancer and duration of follow-up.

**RESULTS:** Twenty-six studies involving more than 2223,704 patients with PC and more than 86034 cases of SMs were entered into this study. The meta-analysis showed that the risk of cancer after PC was 1.03 (95% CI 0.90 - 1.15) and the SIRs of some cancers such as the bladder 1.52 (1.06 - 1.99) and melanoma 1.32 (0.78 - 1.87) were higher than expected. While, malignancies such as rectum 0.92 (0.85 - 1.00), lung 0.85 (0.74 - 0.96) and liver 0.76 (0.54 - 0.98) showed lower incidence in compare to general population.

**CONCLUSIONS AND RELEVANCE:** The overall risk of SMs in patients with prostate cancer is not significantly different from general population, and even in patients undergoing prostatectomy or brachytherapy, the risk is lower. But the incidence of some cancers such as melanoma, bladder, and urinary tract appears to be higher than the public in all types of treatment approaches.

**Key Points:** *Questio:* Is the risk of secondary malignancy in patients with prostate cancer higher than the general population?

*Findings:* This systematic review and meta-analysis of 26 unique trials including 2223,704 patients, showed that the SIRs of some cancers such as the bladder and melanoma were higher than expected.

*Meaning:* These findings suggest that the overall risk of some cancer such as bladder and melanoma in patients with prostate cancer were higher than the general population.

## 1. Introduction

Prostate cancer is the second common cancer among males worldwide. In the United States, it is also the most prevalent non-cutaneous cancer in this sex group, and the second leading cause of death, however, evidence suggests good survival rate in many patients [1]. Factors such as old age, late diagnosis and unsuccessful treatments may lead patients to be at higher risk of death [2]. In a small number of patients, who suffer from advanced stages of cancer, treatment options such as prostatectomy, radiotherapy and more commonly, androgen-deprivation therapy (ADT) have been considered [3]. These approches are associated with different side effects such as urinary incontinence and erectile dysfunction [4, 5]. There are some treatment side effects that could result in hospitalization, including urinary system, reproductive system and gastrointestinal tract disorders [6-8]. Secondary malignancies are one of the complications that could be caused by radiation therapy in cancer patients. Several studies have shown an increased risk of secondary malignancies in patients with prostate cancer [9, 10].

In a study, Lee *et al*. assessed the risk of radiation therapy-induced secondary malignancies in patients with prostate and rectal cancer [11]. Also, in a meta-analysis study, Jin *et al*. examined the incidence of secondary malignancies in patients with prostate cancer following radiotherapy [12].

Herein, we carried out a systematic review and meta-analysis to investigate the incidence of secondary malignancies in patients with prostate cancer in compare to the general population.

## 2. Materials and Methods

### 2.1. Source information

In this systematic review and meta-analysis, we have searched for longitudinal studies through databases of Web of Science, Scopus and PubMed for all available data up to September 2019.

### 2.2. Search strategy

The search was done in the mentioned databases using the following keywords: prostate neoplasm, incidence, second, secondary, after and neoplasms in the title and abstract fields. To do the manual search, the reference list of all eligible papers was also reviewed (Supplement Tabble 1).

### 2.3. Eligibility Criteria

The following inclusion and exclusion criteria were used to select the papers:

- Studies with longitudinal design on prostate cancer patients.
- Studies that declared the results in SIR or those that the SIR could be calculated.
- Articles in English or the available English version published before September 2019.
- No limit was placed on the location of studies.
- All non-longitudinal studies including case series, case-control, and case reports were eliminated.
- All the animal and laboratorial studies were excluded.

### 2.4. Study Selection

First, duplicate studies were deleted by EndNote software, then two researchers (RA and KH) screened the papers independently. We questioned a third researcher as a referee about any disagreement between the two primary researchers. In this meta-analysis risk assessment studies of second malignancies in prostate cancer patients were used.

### 2.5. Quality assessment

For quality assessment, the Newcastle-Ottawa Quality Assessment Form for Cohort Studies (NOS) was used [13]. The studies were classified into three categories; poor (scores 1-3), fair (scores 4-6), and good quality studies (scores 7-9).

### 2.6. Data Extraction

Full texts were reviewed and information such as the author name, year of publication, average age of the patients, SIR of secondary malignancy, and the length of the follow up was recorded.

### 2.7. Statistical Analysis

SIR has calculated by dividing the observed case to the expected case (O/E). Observe means the number of detected cases in the population. And the expected variant is calculated by the age-specific variant of the normal population. SIR helps the researchers to compare and evaluate the rate of incidents to the normal population.

The heterogeneity was evaluated using the *I*^*2*^ test. According to the results and in case of *I*^*2*^ ≥ 50%, the random effect model was used to combine the results. To identify the cause of heterogeneity in the studies, the analysis of sub-groups was performed based on the site of secondary malignancy, the treatment procedure, and duration of follow-up. Data were analyzed using STATA version 11.

### 2.8 Ethical approval

The proposal of this systematic review and meta-analysis has been registered in research committee of Mazandaran University of Medical Sciences with the code number: 6562

## 3. Results

### 3.1. Study selection process

A total of 2210 studies were found in Scopus, PubMed, and Web of Science databases. After excluding duplicate papers, 937 studies remained, which entered the screening stage. Based on the title, abstract and inclusion/exclusion criteria, 58 studies were screened. Finally, 23 studies were selected after reviewing the full texts. After conducting a manual search of the sources, three other studies were found. Eventually, 26 studies that met the inclusion criteria were entered into meta-analysis. (Fig. 1)

**Figure 1.**
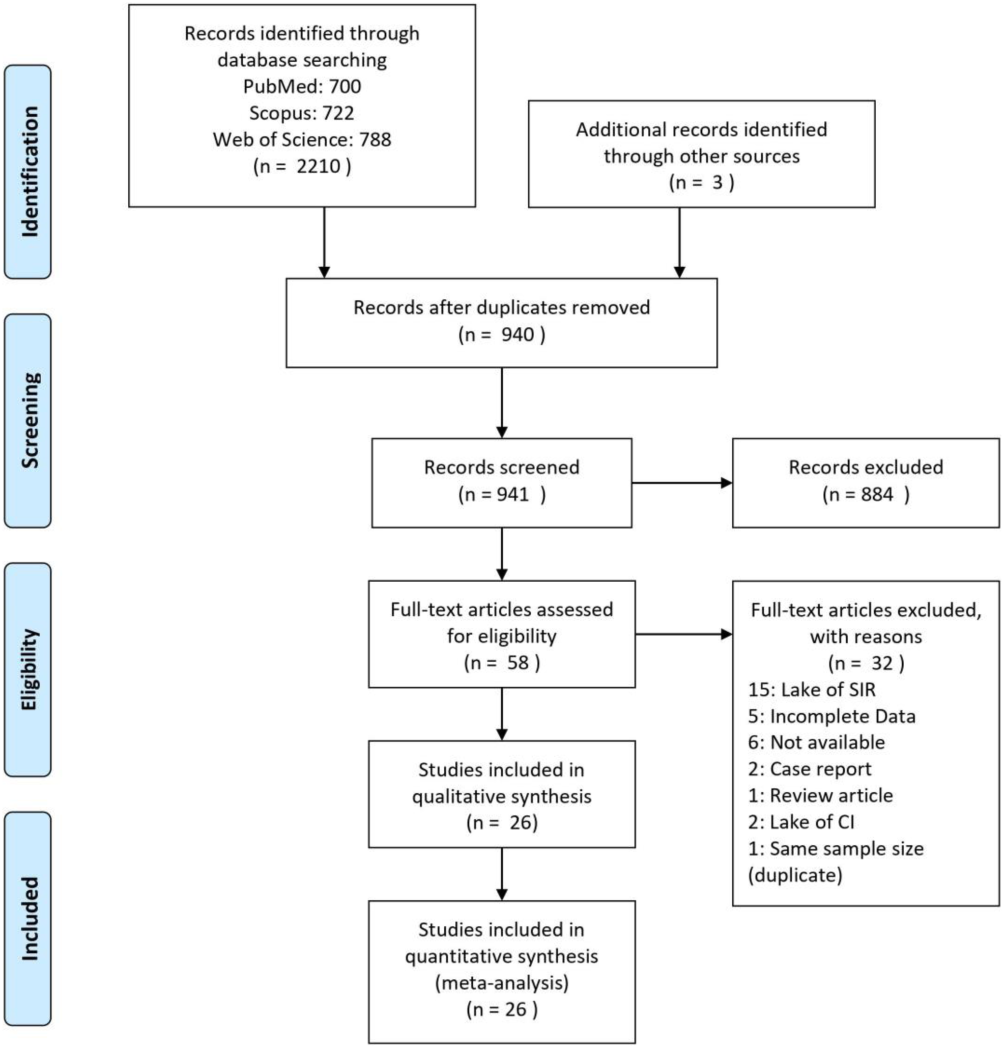
PRISMA flowchart for study selection process.

### 3.2. Study characteristics

The total number of patients was 2,223,704 (406 - 635910). Over 86034 cases of secondary malignancy were observed after the diagnosis and treatment of prostate cancer. The cohort studies were done in the United States (n=7), Switzerland (n= 3), France (n= 3), Sweden (n=2), Australia (n=2), Netherlands (n=1), Israel (n=1), Canada (n=1), Egypt (n=1), Korea (n=1), Norway (n=1), UK (n=1), and Japan (n= 1). One study did not clearly report the location of study. The characteristics of the studies included in the meta-analysis are presented in Table 1.

**Table 1.** Characteristics of studies entered into the meta-analysis

| Author (Year) | Region | Follow-up | Person year Follow-up | Number of patient | Number of Secondary Malignancy | Total Mean Age (range) | Type of Treatment | QA Score |
| --- | --- | --- | --- | --- | --- | --- | --- | --- |
| McCredie et al. (1996) [35] | Australia | 3.4 Y | 77341 | 24163 | 140 | 73.0 | - | 6 |
| Levi et al. (1999) [33] | Switzerland | - | 17065 | 4503 | 380 | 74.2 (42 -97) | - | 5 |
| Thellenberg et al. (2003) [38] | Sweden | 6 Mo | 311996 | 135713 | 10526 | - | - | 4 |
| Rapiti et al. (2008) [37] | Switzerland | 7.4Y | 3797 | 1134 | 19 | 70 (44 – 93) | RT | 6 |
| Nieder et al. (2008) [36] | USA | 49 Mo | 2541930 | 243082 | 3066 | - | RP, RT, BT, RT+BT | 6 |
| Cluze et al. (2009) [14] | France | 3.5 Y | 12601.4 | 3746 | 241 | 72.5 | - | 7 |
| Huo et al. (2019) [29] | USA | - | 3420432 | 635910 | 10179 | 66.9 ± 9.6 | RT, No RT | 5 |
| Zhang et al. (2009) [39] | Sweden | - | - | 18207 | 560 | - | - | 5 |
| Hinnen et al.(2011) [18] | Netherland | 7.5 Y | 14380 | 1888 | 223 | 64.7 | BT, RP | 8 |
| Margel et al. (2011) [34] | Israel | 11.2 Y | - | 29593 | 194 | 70 | RP, RT | 6 |
| Zelevsky et al. (2012) [25] | - | NC | - | 1310 | 115 | NC | RT, BT | 7 |
| Hamilton et al. (2014) [17] | Canada | NC | - | 6433 | 477 | - | RP, BT | 7 |
| Davis et al. (2014) [16] | USA | NC | - | 441504 | 44310 | - | RT, No RT | 7 |
| Van-Hemelrijck et al. (2014) [24] | Switzerland | - | - | 20559 | 1718 | 71.65 ± 9.20 | RT, ADT, RP | 8 |
| Musunuru et al. (2014) [20] | UK | 8 Y | - | 1574 | 194 | 63 (58 – 68) | BT | 7 |
| Joung et al. (2015) [31] | Korea | 42 Mo | - | 55378 | 2761 | 70 | RT, ADT, RP, CT | 6 |
| Keehn et al. (2016) [32] | USA | - | 1973035.5 | 346429 | 6401 | 71 (66 – 75) | BT, RT, | 5 |
| Fernandez-Ots et al. (2016) [21] | Australia | 4.16 | 4104 | 889 | 60 | 63 (40 – 89) | BT | 7 |
| Shahait et al. (2017) [23] | USA | 108.9 Mo | - | 406 | 41 | 62 | - | 7 |
| Abdel-Rahman (2017) [26] | USA | NC | - | - | 3179 | - | - | 4 |
| Cosset et al. (2017) [15] | France | 132 Mo | - | 675 | 61 | 64.5 | BT | 7 |
| Jegu et al. (2017) [30] | France | - | - | NC | NC | - | - | 5 |
| Eweidah et al. (2018) [28] | Egypt | - | - | 152086 | 86 | - | NC | 5 |
| Saltus et al. (2019) [22] | USA | 7 Y | 2922 | 2234 | 172 | 76.6 | RP, No RP | 7 |
| Mohamad et al. (2019) [19] | Japan | 7.9 | 63076.5 | 38594 | 931 | 68 (63 – 73) | BT, RT, RP | 8 |
| Aksnessaether et al. (2019) [27] | Norway | 5.50 Y | 358635 | 57694 | NC | 70 | RT, BT, RT+RP, BT | 6 |

RT: Radiotherapy, RP: Radical Prostatectomy, BT: Brachytherapy, ADT: Androgen Deprivation therapy, CT: Chemotherapy, Y: Year, Mo: Month, Not clear, QA: Quality Assessment

### 3.3. Quality assessment

According to the Newcastle-Ottawa Quality Assessment Form for Cohort Studies, 12 studies had good qualities [14-25] and 14 studies have categorized in fair quality [26-39]. (Supplement Fig. 80 and 82)

### 3.4. Risk of secondary malignancies

According to the investigated studies, 41 different types of secondary malignancies have been reported for prostate cancer patients.

Malignancies that were reported in only one study and were rare included bone and soft tissue-related malignancies [16], connective tissue [31], oral cavity (Buccal) [31], lip [24], respiratory system [31] and ureter [31], Hodgkin’s lymphoma [16], Sarcoma [27], and genital cancers [31].

Malignancies that were observed in more than one study included mouth/pharynx, Esophagus, stomach, small intestine, colon, rectum, anus, colorectal, liver, gallbladder, pancreas, digestive system, larynx, lung, breast, prostate (in patients who received radiotherapy or partial prostatectomy), testis, head and neck cancer, kidney, bladder, urinary tract, skin (melanoma), brain and central nervous system, thyroid gland, endocrine glands, lymphoma, multiple myeloma, leukemia and ultimately hematopoietic cancers.

Secondary malignancies reported in more than one study entered into the current meta-analysis.

In one study, the incidence of SM in patients with prostate cancer treated with ADT was also investigated, but due to paucity of similar information, we could not perform subgroup analysis [24].

SIR of all types of cancers was obtained in three cohort studies from the combination of the risk of other cancers. The SIR of colorectal cancer was found in five studies, and calculated from the combination of SIR of colon cancer and SIR of rectal cancer. SIR of the gastrointestinal tract, urinary tract, and hematopoietic cancers were available in 10, nine, and four studies, respectively, and calculated from the combination of other values reported in these studies.

#### 3.4.1. All malignancies

Overall SIR of secondary malignancies in patients who experienced multiple therapies was 1.03 (95% CI 0.90 – 1.15) (Fig. 2).

**Figure 2.**
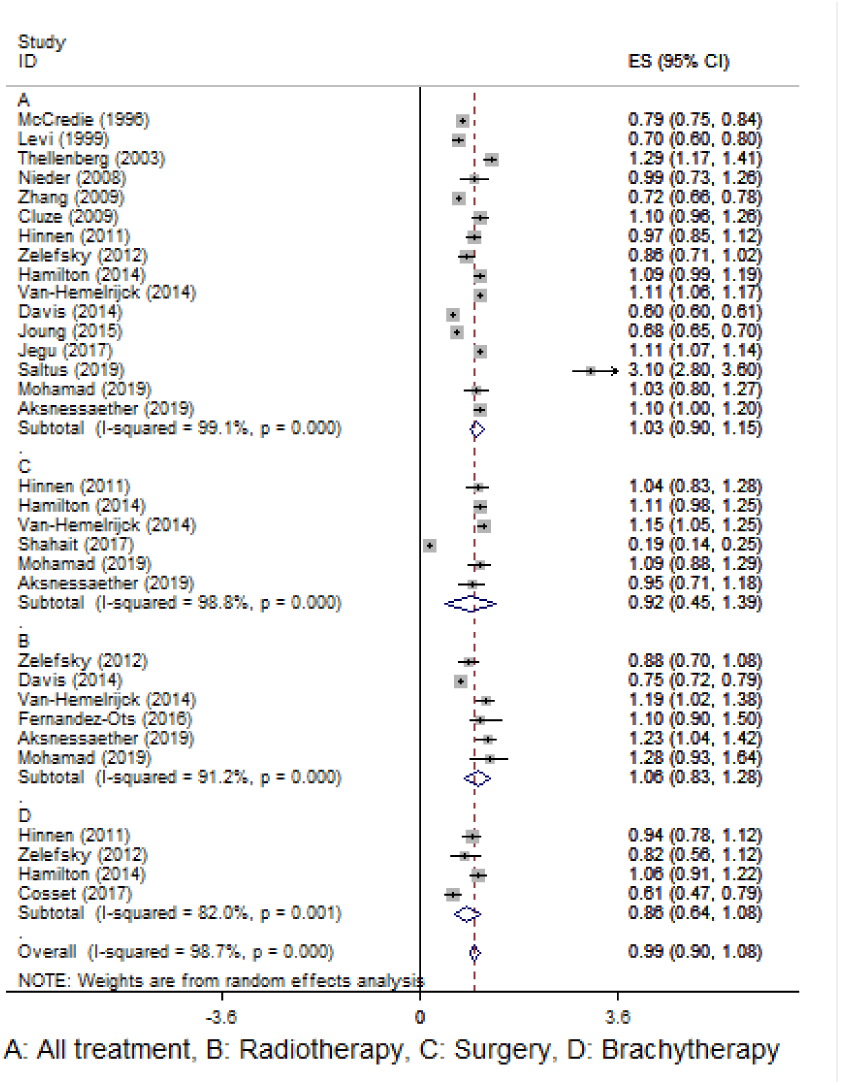
Forest plot for pooling SIR of all cancer.

Overall incidence of secondary malignancies in patients receiving only surgery, radiotherapy, or brachytherapy was 0.92 (95% CI 0.45 – 1.39), 1.06 (95% CI 0.83 – 1.28) and 0.86 (95% CI 0.64 – 1.08), respectively (Fig. 2).

#### 3.4.2. Site of secondary malignancies

##### 3.4.2.1. Digestive system

Total SIR of Mouth and pharynx malignancies was 0.76 (95% CI 0.72 - 0.80) (Supplement Fig. 9 and Table 2).

**Table 2.**
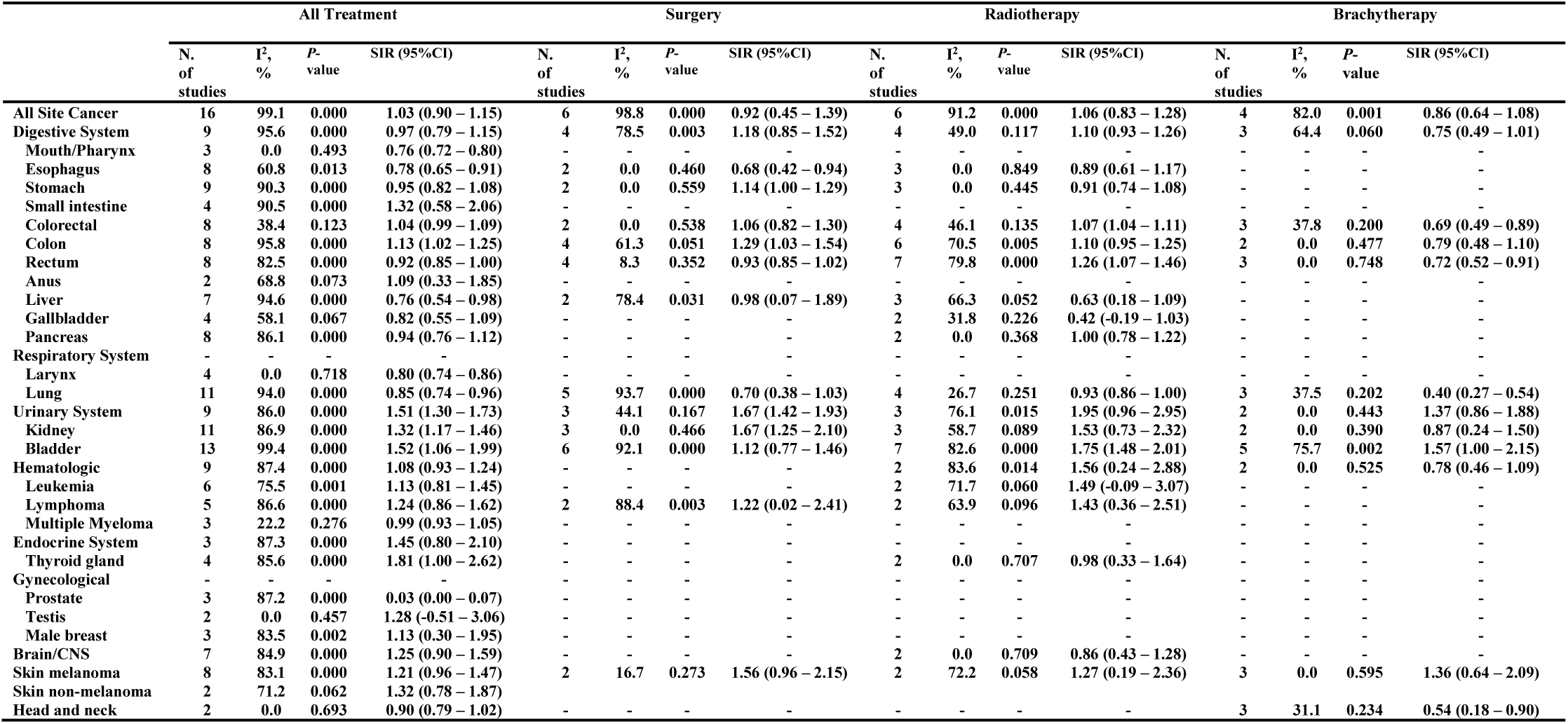
Summarized Pooled SIRs of Considered secondary malignancies

|  | All Treatment |  |  |  | Surgery |  |  |  | Radiotherapy |  |  |  | Brachytherapy |  |  |  |
| --- | --- | --- | --- | --- | --- | --- | --- | --- | --- | --- | --- | --- | --- | --- | --- | --- |
|  | N.<br>of<br>studies | I<sup>2</sup>,<br>% | P-value | SIR (95%CI) | N.<br>of<br>studies | I<sup>2</sup>,<br>% | P-value | SIR (95%CI) | N.<br>of<br>studies | I<sup>2</sup>,<br>% | P-value | SIR (95%CI) | N.<br>of<br>studies | I<sup>2</sup>,<br>% | P-value | SIR (95%CI) |
| All Site Cancer | 16 | 99.1 | 0.000 | 1.03 (0.90 – 1.15) | 6 | 98.8 | 0.000 | 0.92 (0.45 – 1.39) | 6 | 91.2 | 0.000 | 1.06 (0.83 – 1.28) | 4 | 82.0 | 0.001 | 0.86 (0.64 – 1.08) |
| Digestive System | 9 | 95.6 | 0.000 | 0.97 (0.79 – 1.15) | 4 | 78.5 | 0.003 | 1.18 (0.85 – 1.52) | 4 | 49.0 | 0.117 | 1.10 (0.93 – 1.26) | 3 | 64.4 | 0.060 | 0.75 (0.49 – 1.01) |
| Mouth/Pharynx | 3 | 0.0 | 0.493 | 0.76 (0.72 – 0.80) | - | - | - | - | - | - | - | - | - | - | - | - |
| Esophagus | 8 | 60.8 | 0.013 | 0.78 (0.65 – 0.91) | 2 | 0.0 | 0.460 | 0.68 (0.42 – 0.94) | 3 | 0.0 | 0.849 | 0.89 (0.61 – 1.17) | - | - | - | - |
| Stomach | 9 | 90.3 | 0.000 | 0.95 (0.82 – 1.08) | 2 | 0.0 | 0.559 | 1.14 (1.00 – 1.29) | 3 | 0.0 | 0.445 | 0.91 (0.74 – 1.08) | - | - | - | - |
| Small intestine | 4 | 90.5 | 0.000 | 1.32 (0.58 – 2.06) | - | - | - | - | - | - | - | - | - | - | - | - |
| Colorectal | 8 | 38.4 | 0.123 | 1.04 (0.99 – 1.09) | 2 | 0.0 | 0.538 | 1.06 (0.82 – 1.30) | 4 | 46.1 | 0.135 | 1.07 (1.04 – 1.11) | 3 | 37.8 | 0.200 | 0.69 (0.49 – 0.89) |
| Colon | 8 | 95.8 | 0.000 | 1.13 (1.02 – 1.25) | 4 | 61.3 | 0.051 | 1.29 (1.03 – 1.54) | 6 | 70.5 | 0.005 | 1.10 (0.95 – 1.25) | 2 | 0.0 | 0.477 | 0.79 (0.48 – 1.10) |
| Rectum | 8 | 82.5 | 0.000 | 0.92 (0.85 – 1.00) | 4 | 8.3 | 0.352 | 0.93 (0.85 – 1.02) | 7 | 79.8 | 0.000 | 1.26 (1.07 – 1.46) | 3 | 0.0 | 0.748 | 0.72 (0.52 – 0.91) |
| Anus | 2 | 68.8 | 0.073 | 1.09 (0.33 – 1.85) | - | - | - | - | - | - | - | - | - | - | - | - |
| Liver | 7 | 94.6 | 0.000 | 0.76 (0.54 – 0.98) | 2 | 78.4 | 0.031 | 0.98 (0.07 – 1.89) | 3 | 66.3 | 0.052 | 0.63 (0.18 – 1.09) | - | - | - | - |
| Gallbladder | 4 | 58.1 | 0.067 | 0.82 (0.55 – 1.09) | - | - | - | - | 2 | 31.8 | 0.226 | 0.42 (-0.19 – 1.03) | - | - | - | - |
| Pancreas | 8 | 86.1 | 0.000 | 0.94 (0.76 – 1.12) | - | - | - | - | 2 | 0.0 | 0.368 | 1.00 (0.78 – 1.22) | - | - | - | - |
| Respiratory System | - | - | - | - | - | - | - | - | - | - | - | - | - | - | - | - |
| Larynx | 4 | 0.0 | 0.718 | 0.80 (0.74 – 0.86) | - | - | - | - | - | - | - | - | - | - | - | - |
| Lung | 11 | 94.0 | 0.000 | 0.85 (0.74 – 0.96) | 5 | 93.7 | 0.000 | 0.70 (0.38 – 1.03) | 4 | 26.7 | 0.251 | 0.93 (0.86 – 1.00) | 3 | 37.5 | 0.202 | 0.40 (0.27 – 0.54) |
| Urinary System | 9 | 86.0 | 0.000 | 1.51 (1.30 – 1.73) | 3 | 44.1 | 0.167 | 1.67 (1.42 – 1.93) | 3 | 76.1 | 0.015 | 1.95 (0.96 – 2.95) | 2 | 0.0 | 0.443 | 1.37 (0.86 – 1.88) |
| Kidney | 11 | 86.9 | 0.000 | 1.32 (1.17 – 1.46) | 3 | 0.0 | 0.466 | 1.67 (1.25 – 2.10) | 3 | 58.7 | 0.089 | 1.53 (0.73 – 2.32) | 2 | 0.0 | 0.390 | 0.87 (0.24 – 1.50) |
| Bladder | 13 | 99.4 | 0.000 | 1.52 (1.06 – 1.99) | 6 | 92.1 | 0.000 | 1.12 (0.77 – 1.46) | 7 | 82.6 | 0.000 | 1.75 (1.48 – 2.01) | 5 | 75.7 | 0.002 | 1.57 (1.00 – 2.15) |
| Hematologic | 9 | 87.4 | 0.000 | 1.08 (0.93 – 1.24) | - | - | - | - | 2 | 83.6 | 0.014 | 1.56 (0.24 – 2.88) | 2 | 0.0 | 0.525 | 0.78 (0.46 – 1.09) |
| Leukemia | 6 | 75.5 | 0.001 | 1.13 (0.81 – 1.45) | - | - | - | - | 2 | 71.7 | 0.060 | 1.49 (-0.09 – 3.07) | - | - | - | - |
| Lymphoma | 5 | 86.6 | 0.000 | 1.24 (0.86 – 1.62) | 2 | 88.4 | 0.003 | 1.22 (0.02 – 2.41) | 2 | 63.9 | 0.096 | 1.43 (0.36 – 2.51) | - | - | - | - |
| Multiple Myeloma | 3 | 22.2 | 0.276 | 0.99 (0.93 – 1.05) | - | - | - | - | - | - | - | - | - | - | - | - |
| Endocrine System | 3 | 87.3 | 0.000 | 1.45 (0.80 – 2.10) | - | - | - | - | - | - | - | - | - | - | - | - |
| Thyroid gland | 4 | 85.6 | 0.000 | 1.81 (1.00 – 2.62) | - | - | - | - | 2 | 0.0 | 0.707 | 0.98 (0.33 – 1.64) | - | - | - | - |
| Gynecological | - | - | - | - | - | - | - | - | - | - | - | - | - | - | - | - |
| Prostate | 3 | 87.2 | 0.000 | 0.03 (0.00 – 0.07) | - | - | - | - | - | - | - | - | - | - | - | - |
| Testis | 2 | 0.0 | 0.457 | 1.28 (-0.51 – 3.06) | - | - | - | - | - | - | - | - | - | - | - | - |
| Male breast | 3 | 83.5 | 0.002 | 1.13 (0.30 – 1.95) | - | - | - | - | - | - | - | - | - | - | - | - |
| Brain/CNS | 7 | 84.9 | 0.000 | 1.25 (0.90 – 1.59) | - | - | - | - | 2 | 0.0 | 0.709 | 0.86 (0.43 – 1.28) | - | - | - | - |
| Skin melanoma | 8 | 83.1 | 0.000 | 1.21 (0.96 – 1.47) | 2 | 16.7 | 0.273 | 1.56 (0.96 – 2.15) | 2 | 72.2 | 0.058 | 1.27 (0.19 – 2.36) | 3 | 0.0 | 0.595 | 1.36 (0.64 – 2.09) |
| Skin non-melanoma | 2 | 71.2 | 0.062 | 1.32 (0.78 – 1.87) | - | - | - | - | - | - | - | - | - | - | - | - |
| Head and neck | 2 | 0.0 | 0.693 | 0.90 (0.79 – 1.02) | - | - | - | - | - | - | - | - | 3 | 31.1 | 0.234 | 0.54 (0.18 – 0.90) |

Pooled SIR of esophageal cancers in patients receiving combination therapy, surgical treatment, and radiotherapy was 0.78 (95% CI 0.65 - 0.91), 0.68 (95% CI 0.42 - 0.94) and 0.89 (95% CI 0.61-1.17), respectively (Supplement Fig. 10-12). Stomach cancer in patients receiving multiple therapy, surgical and radiotherapy was 0.95 (95% CI 0.82 – 1.08), 1.14 (95%CI 1.00 – 1.29) and 0.91 (95%CI 0.74 – 1.08) respectively (Supplement Fig. 13-15 and Table 2).

Pooling of SIR for small intestine cancers that occur after prostate cancer in patients receiving combination therapy resulted in 1.32 (95% CI 0.58 - 2.06) (Supplement Fig. 16 and Table 2).

The meta-analysis of SIRs in secondary colorectal cancer in patients with prostate cancer who received multiple therapy, surgical treatment, radiotherapy, and brachytherapy resulted in 1.04 (95% CI 0.99 - 1.09), 1.06 (95% CI 0.82 - 1.30), 1.07 (95% CI 1.04 - 1.11) and 0.69 (05% CI 0.49 0.89), respectively. (Supplement Fig. 17-20 and Table 2).

Pooling of SIR for colon and rectal cancers resulted in 1.13 (95% CI 1.02 - 1.25) and 0.92 (95% CI 0.85 - 1.00), respectively (Supplement Fig. 21 and 25 and Table 2). The meta-analysis of SIRs in colon and rectal cancers incidence in patients treated with radiotherapy resulted in 1.10 (95% CI 0.95 - 1.25) and 1.26 (95% CI 1.07 – 1.46), respectively (Supplement Fig. 23 and 27 and Table 2). The incidence of colon and rectal cancers after surgery showed SIRs of 1.29 (95% CI 1.03 - 1.54) and 0.93 (95% CI 0.85 - 1.02) due to meta-analysis (Supplement Fig. 22 and 26 and Table 2). The meta-analysis of SIRs regarding incidence of colon and rectal cancer in patients treated by brachytherapy was as follows respectively: 0.79 (95% CI 0.48 - 1.10) and 0.72 (95% CI 0.52 - 0.91) (Supplement Fig. 24 and 28 and Table 2).

The incidence of gallbladder cancer was evaluated in patients receiving combination therapy and patients receiving only radiotherapy, of which meta-analysis showed the SIRs of 0.82 (95% CI 0.55 – 1.09) and 0.42 (95%CI -0.19 – 1.03) respectively (Supplement Fig. 33 and 34 and Table 2).

SIR of Liver cancer was evaluated in multiple treatments recipients and patients receiving only radiotherapy. Overall SIR obtained through meta-analysis were 0.76 (95% CI 0.54 – 0.98) and 0.63 (95%CI 0.18 – 1.09), respectively (Supplement Fig. 30-32 and Table 2).

The incidence of pancreatic cancer compared to the general population was evaluated in in patients receiving multiple treatments and patients only undergoing radiotherapy. The meta-analysis of SIRs were as follows: 0.94 (95% CI 0.76 - 1.12) and 1.00 (95% CI 0.78 - 1.22), respectively (Supplement Fig. 35 and 36 and Table 2).

Pooled SIR of Anus cancer in patients who experienced multiple therapies was 1.09 (95%CI 0.33 – 1.85) (Supplement Fig. 29 and Table 2).

##### 3.4.2.2. Urinary system

The overall SIR of Urinary tract cancers were in patients receiving multiple types of treatments, radiotherapy recipients, patients undergoing prostatectomy, and finally patients undergoing brachytherapy was 1.51 (95% CI 1.30 - 1.73), 1.95 (95% CI 0.96 - 2.95), 1.67 (95% CI 1.42 - 1.93) and 1.37 (95% CI 0.86 - 1.88), respectively (Supplement Fig. 42-45 and Table 2).

The combination of SIR of Bladder cancers in patients receiving multiple types of treatment, recipients of radiation therapy, patients undergoing prostatectomy, and patients receiving brachytherapy resulted in 1.52 (95% CI 1.06 - 1.99), 1.75 (95% CI 1.48 - 2.01), 1.12 (95% CI 0.77 - 1.46) and 1.57 (95% CI 1.00 - 2.15), respectively (Fig. 3 and Table 2).

**Figure 3.**
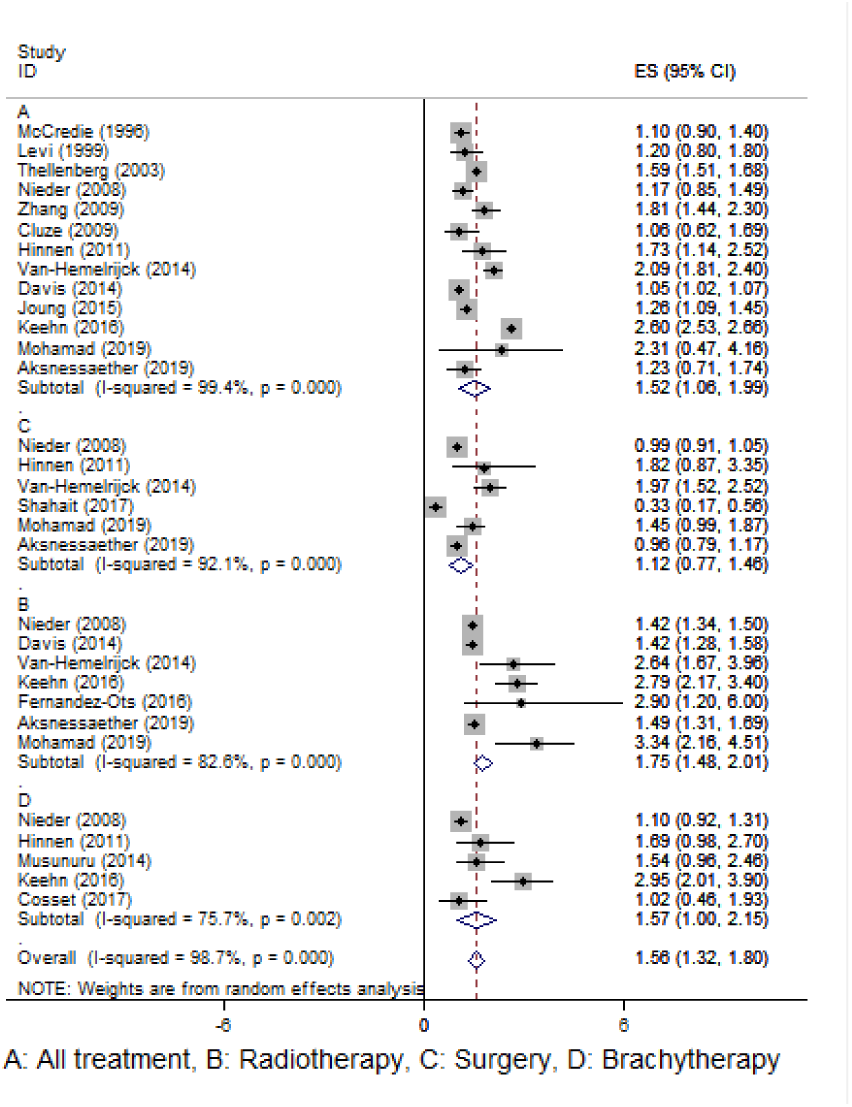
Forest plot for pooling SIR of bladder cancer.

Meta-analysis of kidney cancers in patients receiving multiple treatments, radiotherapy recipients, patients undergoing prostatectomy and brachytherapy resulted in 1.32 (95%CI 1.17 – 1.46), 1.53 (95%CI 0.73 – 2.32), 1.67 (95%CI 1.25 – 2.10) and 0.87 (95%CI 0.24 – 1.50), respectively (Supplement Fig. 46-49 and Table 2).

##### 3.4.2.3. Hematopoietic system

Pooling of SIR for Hematopoietic-related cancers in multi-treatment, radiotherapy and brachytherapy patients resulted in 1.08 (95% CI 0.93 - 1.24), 1.56 (95% CI 0.24 - 2.88) and 0.78 (95% CI 0.46 - 1.09), respectively (Supplement Fig. 54-56 and Table 2).

The overall SIR of leukemia in patients who received several types of treatments and those who received only radiotherapy was 1.13 (95% CI 0.81 - 1.45) and 1.49 (95% CI - 0.09 - 3.07), respectively (Supplement Fig. 57 and 58 and Table 2).

The pooled risk of lymphoma in patients with prostate cancer who received multiple treatments, surgical treatment, and radiotherapy was 1.24 (95% CI 0.86 - 1.62), 1.22 (95% CI 0.02 - 2.41) and 1.43 (95% CI 0.36 - 2.51), respectively (Supplement Fig. 59-61 and Table 2).

The combined SIR of multiple myeloma in patients with prostate cancer who received several types of treatments was 0.99 (95% CI 0.93 - 1.05) (Supplement Fig. 62 and Table 2).

##### 3.4.2.4. Respiratory system

Pooled SIR of lung cancer in patients receiving combination therapy, surgical treatment, radiotherapy, and brachytherapy was 0.85 (95% CI 0.74 - 0.96), 0.70 (95% CI 0.38 - 1.03), 0.93 (95% CI 0.86 - 1.00) and 0.40 (95% CI 0.27 - 0.54), respectively (Supplement Fig. 38-41 and Table 2).

Combination of SIR of Larynx cancer in patients who received multiple types of treatments resulted in 0.80 (95%CI 0.74 – 0.86) (Supplement Fig. 37 and Table 2).

##### 3.4.2.5. Endocrine system

The total SIR of secondary malignancies in the endocrine system in patients who received mixed therapy was 1.45 (95% CI 0.80 - 2.10) (Supplement Fig. 63 and Table 2). Pooled SIR of thyroid malignancies in prostate cancer patients who received multiple therapy and radiotherapy was 1.81 (95%CI 1.00 – 2.62) and 0.98 (95%CI 0.33 – 1.64), respectively (Supplement Fig. 64 and 65 and Table 2).

##### 3.4.2.6. Genital system

Meta-analysis of SIR for the prostate, testis, and breast malignancies resulted in 0.03 (95%CI 0.00 – 0.07), 1.28 (95%CI -0.51 – 3.06), 1.13 (95%CI 0.30 – 1.95), respectively (Supplement Fig. 66- 68 and Table 2).

##### 3.4.2.7. Skin cancers

The summary SIR of melanoma in patients with prostate cancer who received combination therapy, surgical therapy, radiotherapy, and brachytherapy was 1.21 (95%CI 0.96 – 1.47), 1.56 (95%CI 0.96 – 2.15), 1.27 (95%CI 0.19 - 2.36) and 1.36 (95%CI 0.64 – 2.09), respectively (Supplement Fig. 71-74). Meta-analysis of SIR for Non-melanoma skin cancers resulted in 1.32 (95% CI 0.78 - 1.87) (Supplement Fig. 75 and Table 2).

##### 3.4.2.8. Central nervous system

Overall SIR secondary malignancies in the central nervous system in prostate cancer patients who received mixed therapy and radiotherapy was 1.25 (95% CI 0.90 - 1.59) and 0.86 (95% CI 0.43 - 1.28), respectively (Supplement Fig. 69 and 70 and Table 2).

##### 3.4.2.9. Head and neck

Meta-analysis of SIR for head and neck cancer in prostate cancer patients who received multiple therapy and brachytherapy resulted in 0.90 (95% CI 0.79 - 1.02) and 0.54 (95% CI 0.18 - 0.90), respectively (Supplement Fig. 76 and 77 and Table 2).

#### 3.4.3. Duration of follow-up

In this systematic review and meta-analysis, to find the source of heterogeneity in included studies, the SIR of secondary malignancies by the length of follow-ups had been sub-group analyzed.

##### 3.4.3.1. All cancers

In this meta-analysis study, the SIR of all types of solid and non-solid secondary malignancies in patients who received multiple therapy has been evaluated based on the duration of the follow-up period after a prostate cancer diagnosis. Overall SIR in under 5 years of follow up was 0.93 (95% CI 0.63 - 1.22) and in over 5 years of follow up was 0.79 (95% CI 0.64 - 0.94) (Supplement Fig. 80).

##### 3.4.3.2. Bladder cancer

Pooling of SIR of bladder cancer before and after 5 years from prostate cancer diagnosis resulted in 1.15 (95%CI 0.69 – 1.60) and 1.23 (95%CI 0.74 – 1.72), respectively (Supplement Fig. 78).

##### 3.4.3.3. Rectum cancer

Combination of rectum malignancies before and after 5 years from prostate cancer diagnosis resulted in 0.96 (95%CI 0.88 – 1.05) and 1.0 (95%CI 0.74 – 1.29), respectively (Supplement Fig. 79).

## 4. Discussion

We conducted this systematic review and meta-analysis to evaluate the risk of secondary malignancies in patients with prostate cancer. Based on 26 studies entered into meta-analysis, 41 types of secondary malignancies were studied in prostate cancer patients, of which, 31 entered the study. The current study showed that the incidence of digestive system secondary malignancies (stomach, small intestine, colorectal, colon and anus cancers), urinary system (kidney and bladder cancers), hematopoietic system, endocrine system, thyroid gland, testis, breast, central nervous system, cutaneous cancers (melanoma and non-melanoma cancers), leukemia and lymphoma, was higher in prostate cancer patients. At least in one of the subgroups, in contrast to the public. While, some cancers such as oral and laryngeal, esophageal, rectal, liver, gallbladder, pancreas, pharynx, lung, prostate, head and neck, and multiple myeloma malignancies were found to be less frequent than normal population.

One of the major side effects of cancer treatments is cancer relapse and secondary malignancies. It is more important in older patients and in cancers such as testicular in young people. With the development of therapeutic approaches and increased survival rate after receiving cancer treatments, a growing incidence of secondary malignancies is expected.

Given the controversies surrounding the risk of secondary malignancies in patients treated for prostate cancer, to respond the disagreements, we designed a systematic review and meta-analysis and investigated the studies regarding the incidence of secondary malignancies in these patients. The results showed that the risk of secondary malignancies was slightly higher in patients with prostate cancer who received multiple treatments than the general population. Subgroup analysis based on the type of treatment revealed lower risk of cancer in patients undergoing prostatectomy or brachytherapy than those undergoing radiotherapy.

Brachytherapy as a form of local radiotherapy was first introduced by Holm *et al*. [40]. In this treatment, a temporary or permanent source of radiation has inserted into the patient’s prostate glands. The most of the radiations desiring to the malignant tissues and healthy tissues are exposing less radiation. Brachytherapy has become a common treatment option for local, non-metastatic cases in the United States [41]. Given the characteristics of brachytherapy and reduced radiation to healthy tissues compared to radiotherapy, secondary malignancies may be less likely to appear in this type of treatment. In our study, the incidence of secondary malignancies compared to the general population in all cancers except melanoma was lower in patients receiving brachytherapy than in those receiving radiotherapy. Evidence suggests that radiation therapy is more associated with secondary malignancies than brachytherapy [42].

The goal of radiotherapy in patients with prostate cancer is to shed effective amount of radiation that has a tumoricidal effect on the malignant tissue [43]. The volume and anatomy of normal tissues around the prostate should also be considered when performing radiotherapy [44]. The prostate is anatomically located between the rectum and the bladder, so, the normal tissues of these two organs receive the most amount of unnecessary radiation, and there is an increased incidence of secondary malignancies. In patients with prostate cancer, bladder cancer, rectal cancer, and radiation-induced sarcomas are the most common complications following radiotherapy [45]. In current meta-analysis, the incidence of some cancers such as esophageal, stomach, liver, gallbladder, lung, thyroid and central nervous system compared to the normal population, was not only higher, but was also lower. According to some reports, the incidence of rectal cancer in patients with prostate malignancy is not significantly different from the general population [11]. However, the present meta-analysis showed a significant increase in the risk of this type of cancer following radiotherapy. Interestingly, this increase was not observed in the subgroups of patients who received combination therapy, prostatectomy, and brachytherapy. This is in line with the results of Jin *et al*., except that current results had a narrower confidence interval (0.85 – 1.84 and 1.07 – 1.46) [12].

In various studies, the SIR of secondary malignancies in prostate cancer patients was reported separately based on the duration of follow up, indicating lower risk of all cancers compared to that of the general population (at both less and more than 5 years follow up). The findings for rectal cancer were similar to the incidence rates observed in the general population, but observations for bladder cancer showed an increased incidence rate at both follow-up periods. However, the increase was greater at more than 5 years follow-up. Upward trends are reported in the odd ratios over time in the incidence of bladder and rectum cancers [42]. In our meta-analysis, the incidence of all cancers (except rectal and bladder cancers) decreased over time.

The incidence of secondary malignancies after androgen deprivation therapy was analyzed in only one study which did not show an increased rate compared to the normal population [24].

## 5. Conclusions and Implications

Due to the limited number of articles and some disagreements among findings, further research is needed on patients with prostate cancer. Also, the number of studies on the risk of secondary malignancy in patients receiving androgen deprivation therapy is limited. Hence, further investigations on this treatment could be of great benefit in reducing the burden of disease.

According to the results of the present meta-analysis, the overall risk of cancer in patients with prostate cancer is not significantly different from the general population, and even in patients undergoing prostatectomy or brachytherapy, the observed risk is less than expected. But, the incidence of melanoma, bladder, and urinary tract cancers appears to be higher than those in the public after all types of treatments. Nevertheless, the incidence of rectum cancer is higher in patients who receive radiotherapy.

Given the upward trend in the incidence of cancers based on the duration of follow up, younger patients with longer life expectancy should be further evaluated for secondary malignancies.

## Data Availability

The data that support the findings of this study are openly available in data bases mentioned in the search strategy

## 6. Acknowledgment

The authors would like to thank the Student Research Committee of Mazandaran University of Medical Sciences for supporting this project (Project No: 6562).

## Supporting Information Legend

Supplement Figure 1. Forest plot for pooling SIR of All cancer in patient with Multi-treatment

Supplement Figure 2. Forest plot for pooling SIR of All cancer in patient with Prostatectomy

Supplement Figure 3. Forest plot for pooling SIR of All cancer in patient with Radiotherapy

Supplement Figure 4. Forest plot for pooling SIR of All cancer in patient with Brachytherapy

Supplement Figure 5. Forest plot for pooling SIR of Digestive System cancer in patient with Multi-treatment

Supplement Figure 6. Forest plot for pooling SIR of Digestive System cancer in patient with Prostatectomy

Supplement Figure 7. Forest plot for pooling SIR of Digestive System cancer in patient with Radiotherapy

Supplement Figure 8. Forest plot for pooling SIR of Digestive System cancer in patient with Brachytherapy

Supplement Figure 9. Forest plot for pooling SIR of Mouth/Pharynx cancer in patient with Multi-treatment

Supplement Figure 10. Forest plot for pooling SIR of Esophagus cancer in patient with Multi-treatment

Supplement Figure 11. Forest plot for pooling SIR of Esophagus cancer in patient with Prostatectomy

Supplement Figure 12. Forest plot for pooling SIR of Esophagus cancer in patient with Radiotherapy

Supplement Figure 23. Forest plot for pooling SIR of Stomach cancer in patient with Multi-treatment

Supplement Figure 34. Forest plot for pooling SIR of Stomach cancer in patient with Prostatectomy

Supplement Figure 45. Forest plot for pooling SIR of Stomach cancer in patient with Radiotherapy

Supplement Figure 56. Forest plot for pooling SIR of Small intestine cancer in patient with Multi-treatment

Supplement Figure 67. Forest plot for pooling SIR of Colorectal cancer in patient with Multi-treatment

Supplement Figure 78. Forest plot for pooling SIR of Colorectal cancer in patient with Prostatectomy

Supplement Figure 89. Forest plot for pooling SIR of Colorectal cancer in patient with Radiotherapy

Supplement Figure 20. Forest plot for pooling SIR of Colorectal cancer in patient with Brachytherapy

Supplement Figure 21. Forest plot for pooling SIR of Colon cancer in patient with Multi-treatment

Supplement Figure 22. Forest plot for pooling SIR of Colon cancer in patient with Prostatectomy

Supplement Figure 23. Forest plot for pooling SIR of Colon cancer in patient with Radiotherapy

Supplement Figure 24. Forest plot for pooling SIR of Colon cancer in patient with Brachytherapy

Supplement Figure 25. Forest plot for pooling SIR of Rectum cancer in patient with Multi-treatment

Supplement Figure 26. Forest plot for pooling SIR of Rectum cancer in patient with Prostatectomy

Supplement Figure 27. Forest plot for pooling SIR of Rectum cancer in patient with Radiotherapy

Supplement Figure 28. Forest plot for pooling SIR of Rectum cancer in patient with Brachytherapy

Supplement Figure 29. Forest plot for pooling SIR of Anus cancer in patient with Multi-treatment

Supplement Figure 30. Forest plot for pooling SIR of Liver cancer in patient with Multi-treatment

Supplement Figure 31. Forest plot for pooling SIR of Liver cancer in patient with Prostatectomy

Supplement Figure 32. Forest plot for pooling SIR of Liver cancer in patient with Radiotherapy

Supplement Figure 33. Forest plot for pooling SIR of Gallbladder cancer in patient with Multi-treatment

Supplement Figure 34. Forest plot for pooling SIR of Gallbladder cancer in patient with Radiotherapy

Supplement Figure 35. Forest plot for pooling SIR of Pancreas cancer in patient with Multi-treatment

Supplement Figure 36. Forest plot for pooling SIR of Pancreas cancer in patient with Radiotherapy

Supplement Figure 37. Forest plot for pooling SIR of Larynx cancer in patient with Multi-treatment

Supplement Figure 38. Forest plot for pooling SIR of Lung cancer in patient with Multi-treatment

Supplement Figure 39. Forest plot for pooling SIR of Lung cancer in patient with Prostatectomy

Supplement Figure 40. Forest plot for pooling SIR of Lung cancer in patient with Radiotherapy

Supplement Figure 41. Forest plot for pooling SIR of Lung cancer in patient with Brachytherapy

Supplement Figure 42. Forest plot for pooling SIR of Urinary System cancer in patient with Multi-treatment

Supplement Figure 43. Forest plot for pooling SIR of Urinary System cancer in patient with Prostatectomy

Supplement Figure 44. Forest plot for pooling SIR of Urinary System cancer in patient with Radiotherapy

Supplement Figure 45. Forest plot for pooling SIR of Urinary System cancer in patient with Brachytherapy

Supplement Figure 46. Forest plot for pooling SIR of Kidney cancer in patient with Multi-treatment

Supplement Figure 47. Forest plot for pooling SIR of Kidney cancer in patient with Prostatectomy

Supplement Figure 48. Forest plot for pooling SIR of Kidney cancer in patient with Radiotherapy

Supplement Figure 49. Forest plot for pooling SIR of Kidney cancer in patient with Brachytherapy

Supplement Figure 50. Forest plot for pooling SIR of Bladder cancer in patient with Multi-treatment

Supplement Figure 50. Forest plot for pooling SIR of Bladder cancer in patient with Multi-treatment

Supplement Figure 51. Forest plot for pooling SIR of Bladder cancer in patient with Prostatectomy

Supplement Figure 52. Forest plot for pooling SIR of Bladder cancer in patient with Radiotherapy

Supplement Figure 53. Forest plot for pooling SIR of Bladder cancer in patient with Brachytherapy

Supplement Figure 54. Forest plot for pooling SIR of Hematologic cancer in patient with Multi-treatment

Supplement Figure 55. Forest plot for pooling SIR of Hematologic cancer in patient with Radiotherapy

Supplement Figure 56. Forest plot for pooling SIR of Hematologic cancer in patient with Brachytherapy

Supplement Figure 57. Forest plot for pooling SIR of Leukemia cancer in patient with Multi-treatment

Supplement Figure 58. Forest plot for pooling SIR of Leukemia cancer in patient with Radiotherapy

Supplement Figure 59. Forest plot for pooling SIR of Lymphoma cancer in patient with Multi-treatment

Supplement Figure 60. Forest plot for pooling SIR of Lymphoma cancer in patient with Prostatectomy

Supplement Figure 61. Forest plot for pooling SIR of Lymphoma cancer in patient with Radiotherapy

Supplement Figure 62. Forest plot for pooling SIR of Multiple Myeloma cancer in patient with Multi-treatment

Supplement Figure 63. Forest plot for pooling SIR of Endocrine System cancer in patient with Multi-treatment

Supplement Figure 64. Forest plot for pooling SIR of Thyroid gland cancer in patient with Multi-treatment

Supplement Figure 65. Forest plot for pooling SIR of Thyroid gland cancer in patient with Radiotherapy

Supplement Figure 66. Forest plot for pooling SIR of Prostate cancer in patient with Multi-treatment

Supplement Figure 67. Forest plot for pooling SIR of Testis cancer in patient with Multi-treatment

Supplement Figure 68. Forest plot for pooling SIR of Male breast cancer in patient with Multi treatment

Supplement Figure 69. Forest plot for pooling SIR of Brain/CNS cancer in patient with Multi-treatment

Supplement Figure 70. Forest plot for pooling SIR of Brain/CNS cancer in patient with Radiotherapy

Supplement Figure 71. Forest plot for pooling SIR of Skin melanoma cancer in patient with Multi-treatment

Supplement Figure 72. Forest plot for pooling SIR of Skin melanoma cancer in patient with Prostatectomy

Supplement Figure 73. Forest plot for pooling SIR of Skin melanoma cancer in patient with Radiotherapy

Supplement Figure 74. Forest plot for pooling SIR of Skin melanoma cancer in patient with Brachytherapy

Supplement Figure 75. Forest plot for pooling SIR of Skin Non-melanoma cancer in patient with Multi-treatment

Supplement Figure 76. Forest plot for pooling SIR of Head and neck cancer in patient with Multi-treatment

Supplement Figure 77. Forest plot for pooling SIR of Head and neck cancer in patient with Brachytherapy

Supplement Figure 78. Forest plot for pooling SIR of bladder cancer based on follow-up duration

Supplement Figure 79. Forest plot for pooling SIR of rectum cancer based on follow-up duration

Supplement Figure 80. Forest plot for pooling SIR of all cancer based on follow-up duration

Supplement Figure 81. Risk of bias summary (A)

Supplement Figure 82. Risk of bias summary (B)

